# Methamphetamine addiction and its impact on adult clients at the National Institute of Mental Health, Sri Lanka: A descriptive cross-sectional study

**DOI:** 10.1101/2025.06.02.25328838

**Authors:** N.A.A.I. Nishshanka, T.N.L. Samarathunga, S.W. Inoka, R. Suharna, D.K.M. De Silva, K.A. Sriyani

## Abstract

Methamphetamine addiction poses a growing public health challenge in Sri Lanka, yet limited research explores its impacts on the addicted population. This study aimed to assess the severity, patterns, and consequences of Methamphetamine addiction among adult patients at the National Institute of Mental Health (NIMH), Angoda, Sri Lanka. A descriptive cross-sectional study was conducted among adult clients (aged >18 years) diagnosed with Methamphetamine use disorder according to DSM-5 criteria. A sample of 427 participants was recruited through purposive sampling. Data were collected using a structured, validated, interviewer-administered questionnaire covering sociodemographic details, addiction severity, consumption patterns, impacts, and reasons for use. Descriptive statistics were analyzed using SPSS version 26. All participants (100%) responded to the survey. Among participants, 93.7% were male, and 65.3% were aged 18–30 years. The majority resided in urban (57.8%) or semi-urban (36.1%) areas. Addiction severity was categorized as mild (29%), moderate (38.6%), and severe (32.3%). Most (65.3%) initiated Methamphetamine use between 21–30 years. Smoking (52.7%) and snorting (45%) were common methods of use, with peer pressure (48.9%) cited as the primary reason for initiation. The most cited physical impacts were weight loss (38.88%) and loss of appetite (37.24%), while irritability (28.81%) and interpersonal relationship problems (50.82%) were cited as common mental and social impacts, respectively. Findings reveal that young urban males are predominantly affected by Methamphetamine addiction, with moderate to severe dependence common. Peer influence and easy accessibility were significant contributing factors. The physical, mental, and social health impacts emphasize the urgent need for comprehensive intervention strategies focusing on prevention, early detection, and integrated rehabilitation services at the national level.

## Background

Methamphetamine is a potent central nervous system stimulant known for its strong addictive potential, which poses a critical challenge to global public health [1]. The global count of amphetamine users has surged past 27 million in 2020 [2]. According to the 2021 National Survey on Drug Use and Health (NSDUH), More than 16.8 million people age 12 or older used Methamphetamine at least once during their lifetime.

Asian countries also reflected broader trends in Methamphetamine addiction during the last years. Thailand and the Philippines were reported as the highest-rated Methamphetamine prevalence in Asia [3]. In Sri Lanka, the drug abuse landscape is shifting, with Methamphetamine becoming a prevalent concern [4]. In 2022, Methamphetamine was ranked as the third most commonly abused substance in Sri Lanka [5]. This escalation was also evident as, in 2022, 22631 Sri Lankan individuals were arrested related to Methamphetamine offenses, with around 241 kg of Methamphetamine [6]. Notably, in 2023, these values were increased to 26,096 arrested individuals related to Methamphetamine offenses [7]. This report also underscored the gender disparity, with an overwhelmingly male representation, as 99.65% of individuals seeking treatment identified as male [7].

The reasons for Methamphetamine use are complex and multifactorial. For most users, Methamphetamine serves as a tool for enhancing performance, as it can temporarily increase energy, alertness, and concentration, fostering its use among adults engaged in high-pressure environments [8]. Moreover, social factors including peer influences, cultural acceptance, and the accessibility of the drug contribute to its appeal [5]. Several users also reported that past experiences of trauma, mental health issues, and familial drug use histories elevate the likelihood of Methamphetamine use [9]. However, the motivation behind Methamphetamine use was not clear in the Sri Lankan context.

Methamphetamine addiction has widespread effects that are multifaceted and interrelated, which encompass physical, psychological, and social domains [10]. Concerning the physiological impact of chronic Methamphetamine addiction, it leads to significant cardiovascular problems, including increased heart rate and blood pressure, which can culminate in cardiovascular collapse [11]. Other than that, users also reported weight loss, dental issues colloquially known as “meth mouth,” and skin infections due to increased scratching and neglect of personal hygiene [12]. Psychologically, Methamphetamine addiction causes varied mental health issues including immediate euphoria coupled with anxiety, depression, and psychosis [13]. Users usually reported symptoms of cognitive dysfunction, including impaired decision-making and difficulties with impulse control, which are linked to alterations in the prefrontal cortex’s functionality [14]. Moreover, the cycle of withdrawal enhances the psychological burden; users experience a range of withdrawal symptoms, including dysphoria, insomnia, and intense drug cravings, which can perpetuate the cycle of addiction [15].

Moreover, Methamphetamine addiction devastates interpersonal relationships and disrupts family dynamics, leading to heightened social stigma against users and their families [16]. Compulsive behaviors due to Methamphetamine addiction can lead users to engage in criminal activities, high-risk sexual behaviors, which complicates the public health efforts [16,17].

Despite the growing prevalence of Methamphetamine use in Sri Lanka and its evident socio-health impact globally, there remains a critical gap in empirical research, particularly regarding the patterns, motivations, and health consequences of Methamphetamine addiction among adult individuals in Sri Lanka. Most of the available data are based on arrest records and broad treatment statistics, lacking the necessary depth to inform tailored interventions, policy frameworks, or harm-reduction strategies. In light of this, the current study aimed to identify the severity and patterns of Methamphetamine consumption, determine the prevalence and types of polydrug use among MA users, and explore self-reported reasons and impacts of Methamphetamine use in Sri Lanka.

## Method

### Study design and setting

A descriptive cross-sectional study was designed to assess Methamphetamine addiction and its impact on adult patients who were admitted to the National Institute of Mental Health (NIMH) at Angoda, Sri Lanka. NIMH serves as the country’s premier institution for psychiatric and mental health care and addiction management. Patients with mental illness and several drug addictions from all over the country are referred here for treatment. As the largest and only specialized tertiary-level mental health facility in the country, NIMH provides comprehensive mental health services, including inpatient, outpatient, and community-based psychiatric care. It plays a pivotal role in addressing the national burden of mental health disorders, including substance use disorders, and acts as the central referral hub for patients from all parts of the island.

### Population

The study’s target population consisted of all adult individuals (more than 18 years old) receiving treatment for Methamphetamine addiction in the wards and day center at the NIMH during the study period. The decision to include only adults was based on several considerations. Firstly, the clinical presentation, patterns of substance use, psychosocial consequences, and treatment modalities for Methamphetamine addiction differ markedly between adolescents and adults; thus, including only adults ensured a more homogeneous sample and increased the internal validity of the findings. Further, the focus on adult patients aligns with the core patient population of the NIMH’s addiction treatment services, enhancing the study’s relevance to national policy and clinical practice.

Both male and female adult individuals who were diagnosed with Methamphetamine substance abuse disorder according to DSM-5 criteria (F15.10, F15.15, and F15.20) and were currently receiving treatments were included in the current study, while individuals with cognitive impairment and withdrawal symptoms of Methamphetamine were excluded from the study.

### Sample and sample size

The sample size for the study was determined using Daniel’s sample size calculation formula. Assuming a 95% confidence level (Z = 1.96), a prevalence (P) of 50%, and a precision (d) of 0.05, the calculated sample size was 384 [18]. To account for a potential 10% non-response rate (d), the sample size was adjusted using the formula N=n/1−d, resulting in a final required sample size of 427 participants. The list of eligible patients was identified with the support of clinical staff and verified through medical records from wards and day centers. Participants were then recruited using a purposive sampling technique based on the predefined inclusion and exclusion criteria due to the population represents a specific subgroup within the broader psychiatric patient population.

### Data collection tool

A structured, interviewer-administered questionnaire was utilized to collect the data. It was developed by referring to existing literature related to substance abuse and Methamphetamine addiction [19–23] . It was comprised of four sections. The first section of the questionnaire was dedicated to obtaining participants’ socio-demographic data, which had ten items including age, gender, ethnicity, civil status, family status, educational level, occupation, monthly income, residence area, and province of living. The second section assessed the severity of addiction according to DSM-5 criteria[24]. It consisted of 11 items on symptoms of addiction, and the level of severity was categorized as mild, moderate, and severe according to a number of symptoms present with the addicted individual (mild = 2−3, moderate = 4−5, severe = 6−11) [24]. The third section of the questionnaire included nine items to determine the pattern of Methamphetamine consumption, including age of initiation, method of use, frequency of use, accessibility to Methamphetamine, last use, person who introduced Methamphetamine, daily expenditure to use Methamphetamine, and use of other substances (polydrug use). In addition, three open-ended items were used to collect information regarding the physical, psychological, and social impact of MA use reasons for Methamphetamine use. The last section of the questionnaire included self-reported reasons (10 items with ‘yes’ or ‘no’ responses) for Methamphetamine consumption.

The content validity of the questionnaire was ensured with experts’ opinions, including a consultant psychiatrist, a nursing academic, and a trained psychiatric nurse, and necessary modifications were made. The questionnaire was pre-tested among ten patients who were being treated for Methamphetamine addiction to improve its clarity and examine whether the respondents could understand the items they were expected to answer [25]. The questionnaire was finalized considering the participants’ opinions received during the pre-test. Patients who participated in the pre-test were not included in the main study. Reliability was assessed on the 11 items assessing the severity of addiction, indicated acceptable internal consistency, with a Cronbach’s alpha of 0.89 [26].

### Data collection

Data collection was commenced after obtaining ethical clearance for the study from the Ethics Review Committee of NIMH and permission from the relevant hospital authorities. Data were collected from 20^th^ July 2023 to 20^th^ October 2023. All the patients were fully informed about the purpose, nature, risks, and benefits of the study verbally and through an information sheet and obtained written consent before their participation. Volunteer participation was encouraged. Data were collected by four investigators who had undergone a training session on data collection using an interviewer-administered questionnaire. It was done with strict adherence to patient privacy and confidentiality, ensuring that data collection did not interfere with the patient’s treatment or care within their ward setup or daycare center.

## Ethical considerations

Ethical approval for the study was obtained from the Ethics Review Committee of NIMH (ERC No: 205/03/2023). Permission to access the setting and participants was obtained from the Director of the NIMH and Consultant Psychiatrists. Informed consent was signed by all the participants who volunteered for the study. Permission to withdraw from the study was granted to the participants, and anonymity and confidentiality of the participants were ensured.

## Data analysis

The collected data were coded and entered into SPSS version 26 for the analysis. The dataset was cleansed to detect any missing values or outliers. Data were descriptively analyzed for frequencies and percentages. MA addiction was categorized into mild, moderate, and severe based on the DSM-5 established standard framework [23].

## Findings

### Sociodemographic characteristics of the participants

Out of the calculated sample size of 427, all responses yielded a 100% response rate. Table 1 presents the socio-demographic data of the participants. Of the sample, the majority consisted of males (n = 400, 93.68%). Most of the participants fell within the young adult age range of 18-30 years (n = 279, 65.3%). The findings revealed a diverse racial composition, with the Sinhala ethnic group being the most prominent, comprising 59% (n=252) of the total participants.

**Table 1.** Sociodemographic data of the participants (n=427)

| Characteristics | Frequency (n) | Percentage (%) |
| --- | --- | --- |
| **Age (Years)** |  |  |
| 18-30 | 279 | 65.3 |
| 31-40 | 101 | 23.7 |
| 41-50 | 34 | 8 |
| 51-60 | 13 | 3 |
| **Gender** |  |  |
| Male | 400 | 93.7 |
| Female | 27 | 6.3 |
| **Social status** |  |  |
| Married | 150 | 35.1 |
| Single | 183 | 42.9 |
| Widowed | 06 | 1.4 |
| Divorced | 22 | 5.2 |
| Separate | 66 | 15.5 |
| **Education** |  |  |
| Primary education | 63 | 14.8 |
| Secondary education | 254 | 59.5 |
| Tertiary education | 59 | 13.8 |
| Higher education | 06 | 1.4 |
| Vocational education | 45 | 10.5 |
| **Ethnicity** |  |  |
| Sinhala | 252 | 59 |
| Muslim | 105 | 24.6 |
| Tamil | 70 | 16.4 |
| **Living with whom** |  |  |
| Family | 323 | 75.6 |
| Alone | 95 | 22.2 |
| other | 09 | 2.1 |
| **Monthly income (LKR)** |  |  |
| <20,000 | 66 | 15.5 |
| 20,000 – 30,000 | 69 | 16.2 |
| 30,0001 – 40,000 | 136 | 31.9 |
| 40,001 – 50,000 | 90 | 21.1 |
| >50,000 | 66 | 15.5 |
| **Living province** |  |  |
| Western province | 292 | 68.4 |
| Southern province | 29 | 6.8 |
| Sabaragamuwa province | 03 | 0.7 |
| Eastern province | 04 | 0.9 |
| Central province | 21 | 4.9 |
| North Western province | 61 | 14.3 |
| Northern province | 17 | 4.0 |
| **Living area** |  |  |
| Urban | 247 | 57.8 |
| Semi-urban | 154 | 36.1 |
| Rural | 26 | 6.1 |

LKR; Sri Lankan rupees

A total of 42.9%(n=183) of respondents were single. A total of 59.5%(n=254) of individuals attained education up to the secondary level. Most of the participants (n=292, 68.40%) were from the Western Province, and a significant proportion from North Western Province (n=61, 14.30%) followed by Southern Province (n=29, 6.80%). The majority of participants reside in urban (n= 57.8%) and suburban (36.1%) areas in the country.

### MA addiction severity and the pattern of addiction among the participants

The findings of the severity and patterns of Methamphetamine addiction among participants are presented in Table 2. Addiction severity was categorized as mild (29%), moderate (38.6%), and severe (32.3%). The age of onset ranged predominantly between 21-30 years (65.3%), with smaller proportions starting at ages 12-20 (22.5%) and 31-40 (9.8%) years. Consumption frequency varied, with 48.2% using Methamphetamine several days a week, 28.1% consuming daily, and 22.2% using weekly. The majority reported their last usage within a week (54.6%), while 22.7% used it within a month. Smoking (52.7%) and snorting (45%) were the most common methods of consumption, with minimal use of injection (1.9%) or swallowing (0.5%). Most participants were introduced to Methamphetamine by friends (83.8%), followed by relatives (11%). Accessibility to the substance was deemed fairly easy (46.6%) or easy (36.5%) by the majority, while 5.6% found it difficult to access.

**Table 2.**
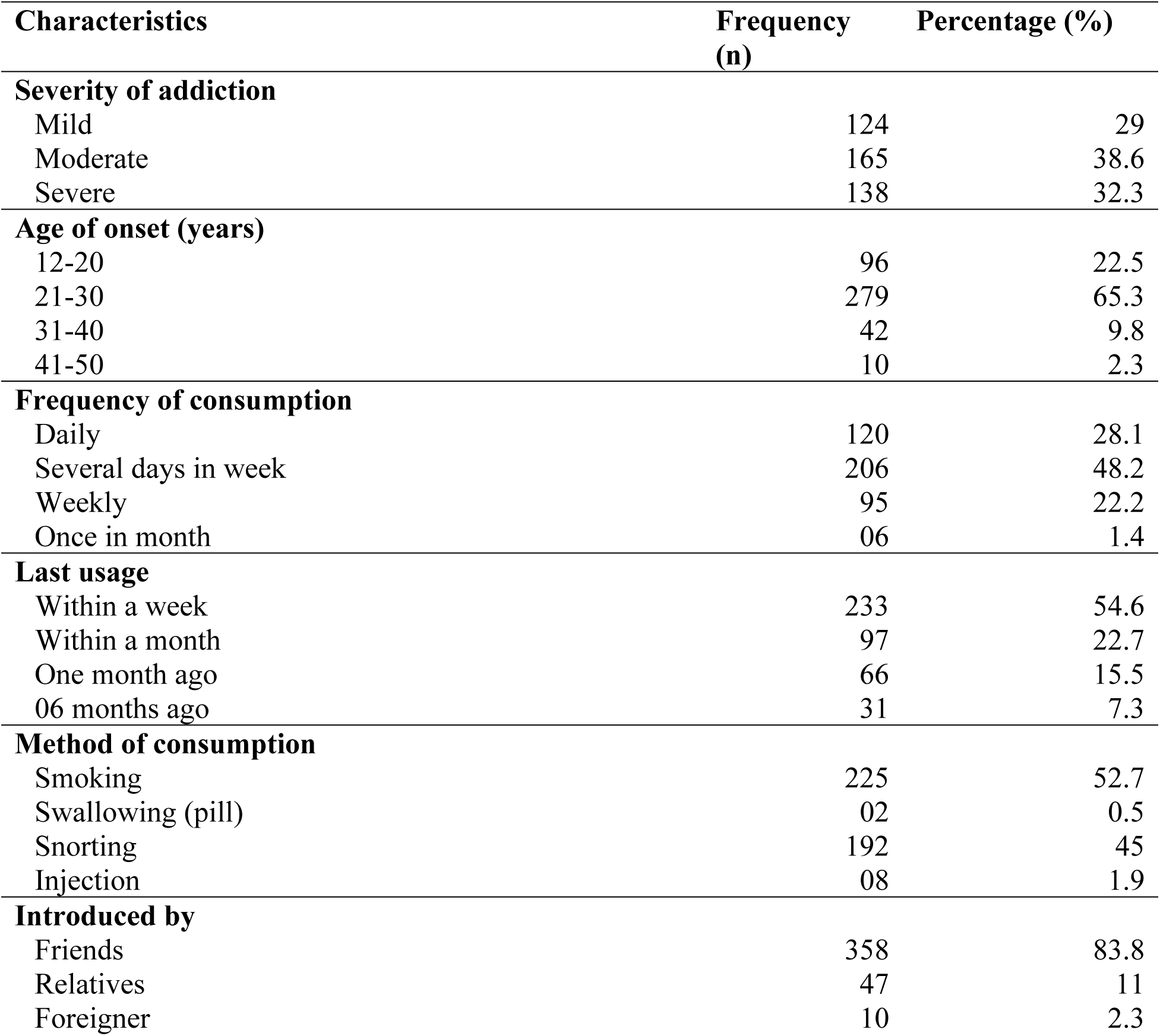
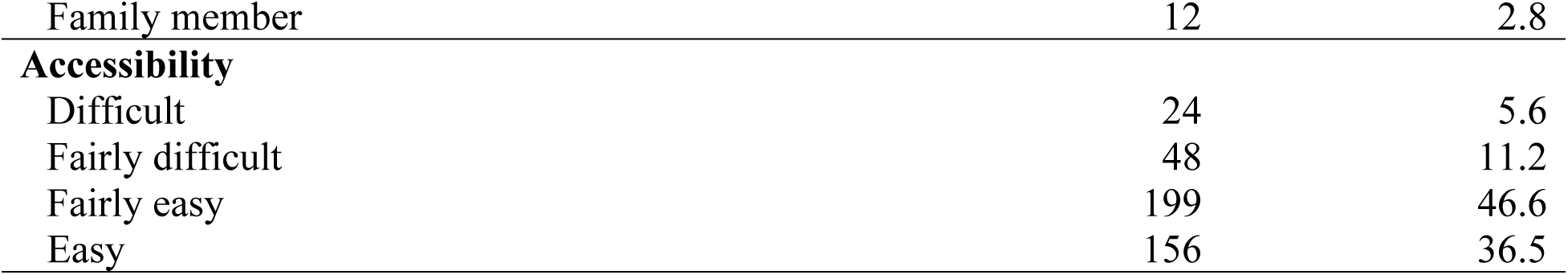
MA addiction severity and the pattern of addiction among the participants

### Polydrug consumption

Out of the sample, 55 (12.9%) consume Methamphetamine alone while the rest of them (n=372, 87.1%) consume other drugs with Methamphetamine. Out of polydrug consumers, 164 (38.4%) are alcohol consumers, 166 (38.9%) use cannabis, 128 (30%) consume heroin, 131 (30.7%) are tobacco consumers, and 111 (26%) consume other drugs.

### Self-reported reasons for MA consumption

Participants were asked to cite the most probable reasons for their Methamphetamine addiction, and individuals were able to cite one or more reasons. Table 3 represents the self-reported reasons for Methamphetamine addiction among the participants. Peer pressure was the most frequently cited reason, with 48.9% acknowledging its influence. A smaller proportion (23.2%) reported the involvement of peers in drug-related businesses as a contributing factor. Family-related reasons, such as isolation from family (11.5%), lack of family closeness (9.1%), and lack of parental support (3.3%), were less commonly reported. Only 1.9% attributed their consumption to having parents who were drug abusers, and 1.2% cited excessive punishment as a reason. Work-related factors included using Methamphetamine to maintain attention and concentration (13.6%), increase job productivity (10.5%), and cope with a heavy workload (2.6%).

**Table 3.**
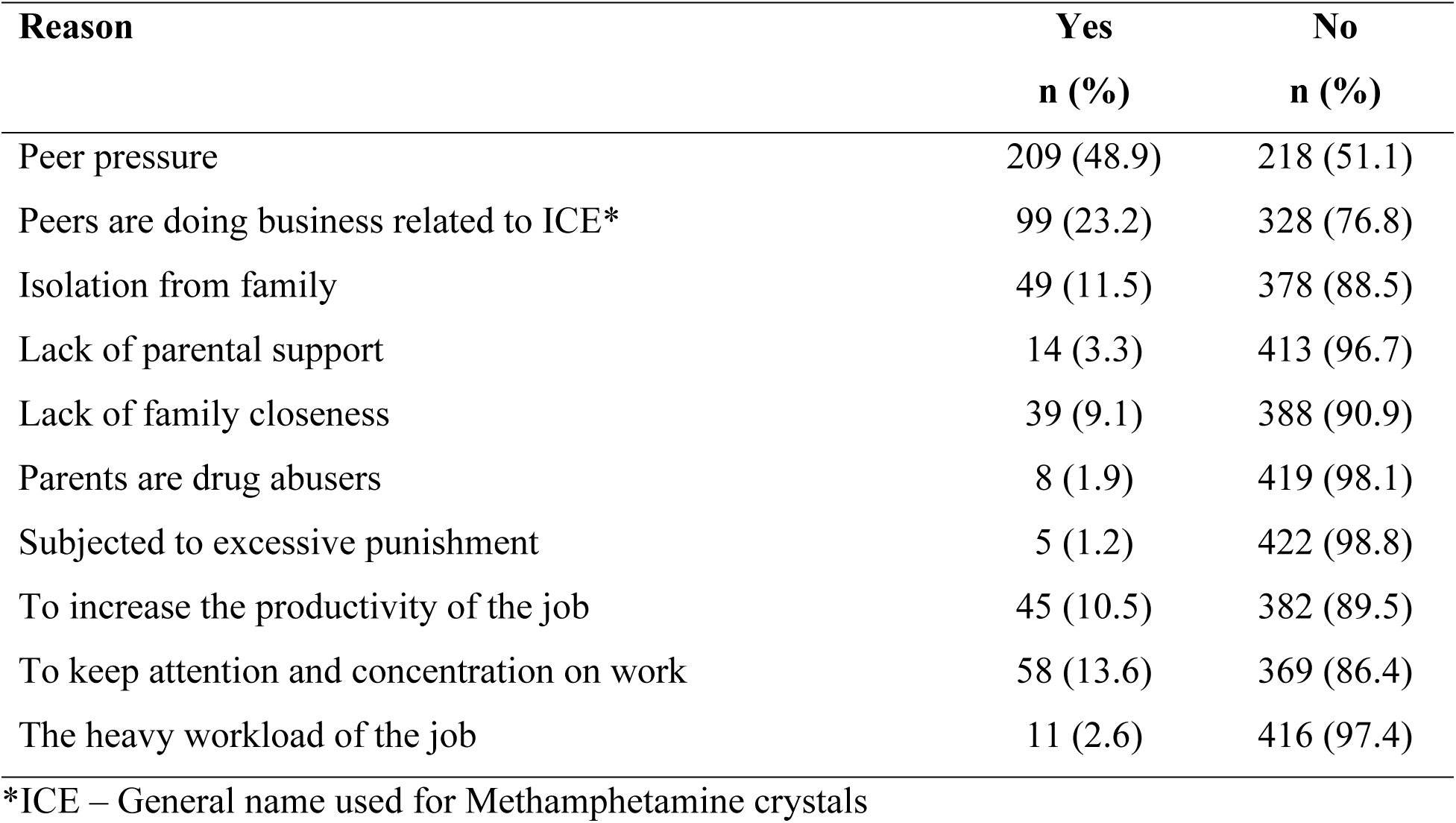
Self-reported reasons for Methamphetamine addiction among the participants.

| Reason | Yes<br>n (%) | No<br>n (%) |
| --- | --- | --- |
| Peer pressure | 209 (48.9) | 218 (51.1) |
| Peers are doing business related to ICE* | 99 (23.2) | 328 (76.8) |
| Isolation from family | 49 (11.5) | 378 (88.5) |
| Lack of parental support | 14 (3.3) | 413 (96.7) |
| Lack of family closeness | 39 (9.1) | 388 (90.9) |
| Parents are drug abusers | 8 (1.9) | 419 (98.1) |
| Subjected to excessive punishment | 5 (1.2) | 422 (98.8) |
| To increase the productivity of the job | 45 (10.5) | 382 (89.5) |
| To keep attention and concentration on work | 58 (13.6) | 369 (86.4) |
| The heavy workload of the job | 11 (2.6) | 416 (97.4) |

\*ICE – General name used for Methamphetamine crystals

### Physical Impact of Methamphetamine

Self-reported physical impacts of Methamphetamine addiction were summarized in Figure 1. Weight loss (38.88%) and loss of appetite (37.24%) were the most commonly reported physical impacts among participants. Dental problems affected 12.18%, while malaise and chest pain were reported by 6.32% each. Other notable impacts included cough (7.49%), dry mouth (5.15%), myalgia (5.62%), and excessive sweating (4.22%). Less frequently reported issues included physical injuries, headaches, and muscle cramps (around 3.28%-3.98% each), hair loss (2.34%), and jaw clenching (1.17%). Muscle rigidity was the least reported, affecting only 0.47% of participants.

**Figure 1.**
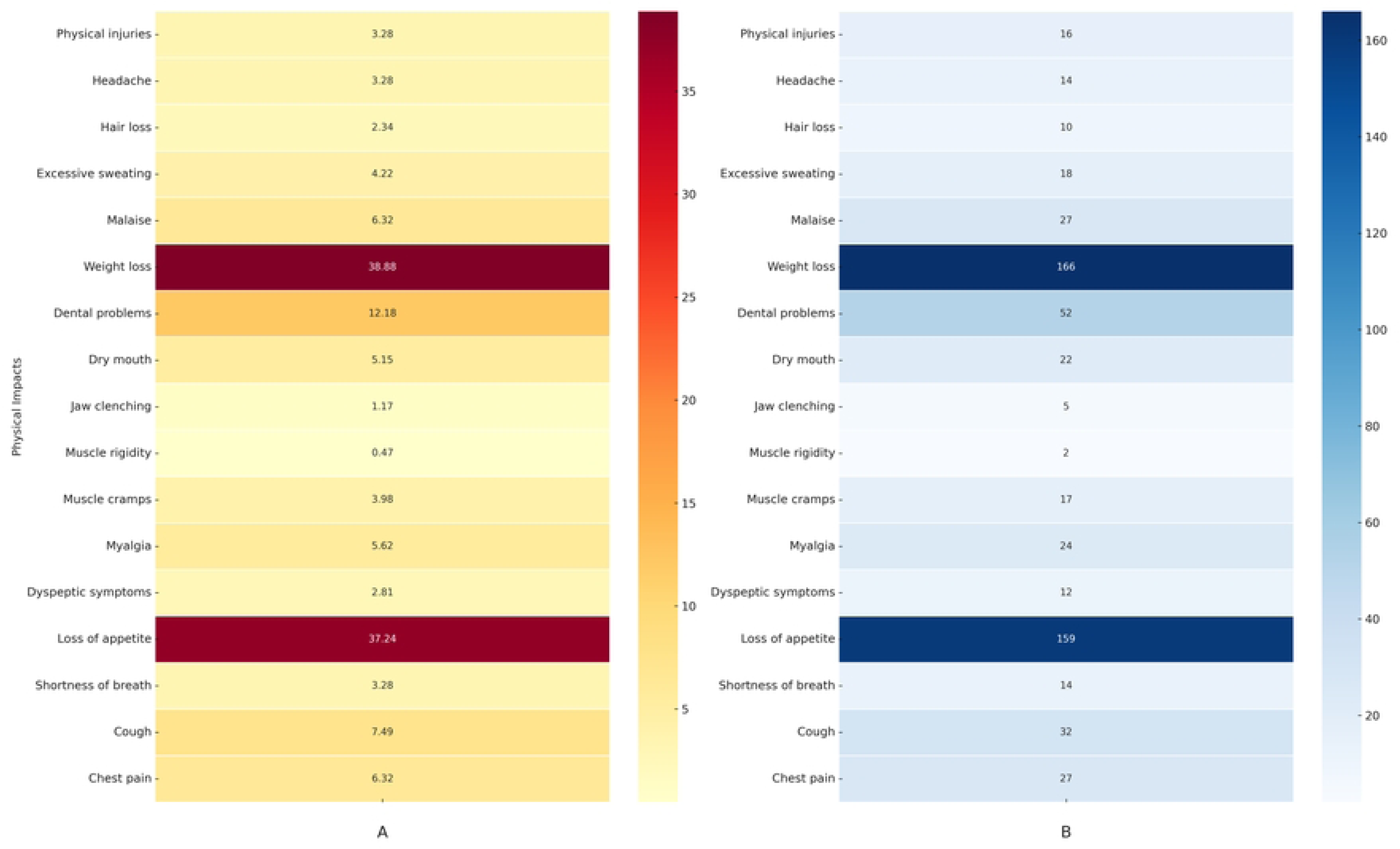
Heatmaps illustrating self-reported physical impacts due to Methamphetamine addiction. *(A) Percentage heatmap: Illustrates the relative percentages of self-reported physical impacts due to Methamphetamine addiction, with darker shades indicating a higher percentage of respondents experiencing each impact*. *(B) Frequency heatmap: Illustrates the absolute number of responders’ self-reported physical impacts due to Methamphetamine addiction, with darker shades indicating a higher number of respondents experiencing each impact*.

### Psychological/mental impact

The psychological impacts of Methamphetamine addiction are shown in Figure 2. Irritability was the most frequently reported psychological symptom (28.81%), followed by delusions (24.82%) and hallucinations (22.95%). Sleep problems were also common, affecting 18.74% of participants. Anxiety and fearfulness (14.52%), feeling low (11.94%), and poor concentration and attention (8.43%) were notable issues. Suicidal thoughts or self-harm behaviors were reported by 7.96%, while homicidal ideas and aggression were less common, affecting 3.04% and 4.68%, respectively. Loss of interest (6.32%) and restlessness/agitation (1.17%) were among the least reported. These findings highlight significant mental health challenges, particularly irritability, psychotic symptoms, and anxiety-related issues, associated with Methamphetamine use.

**Figure 2.**
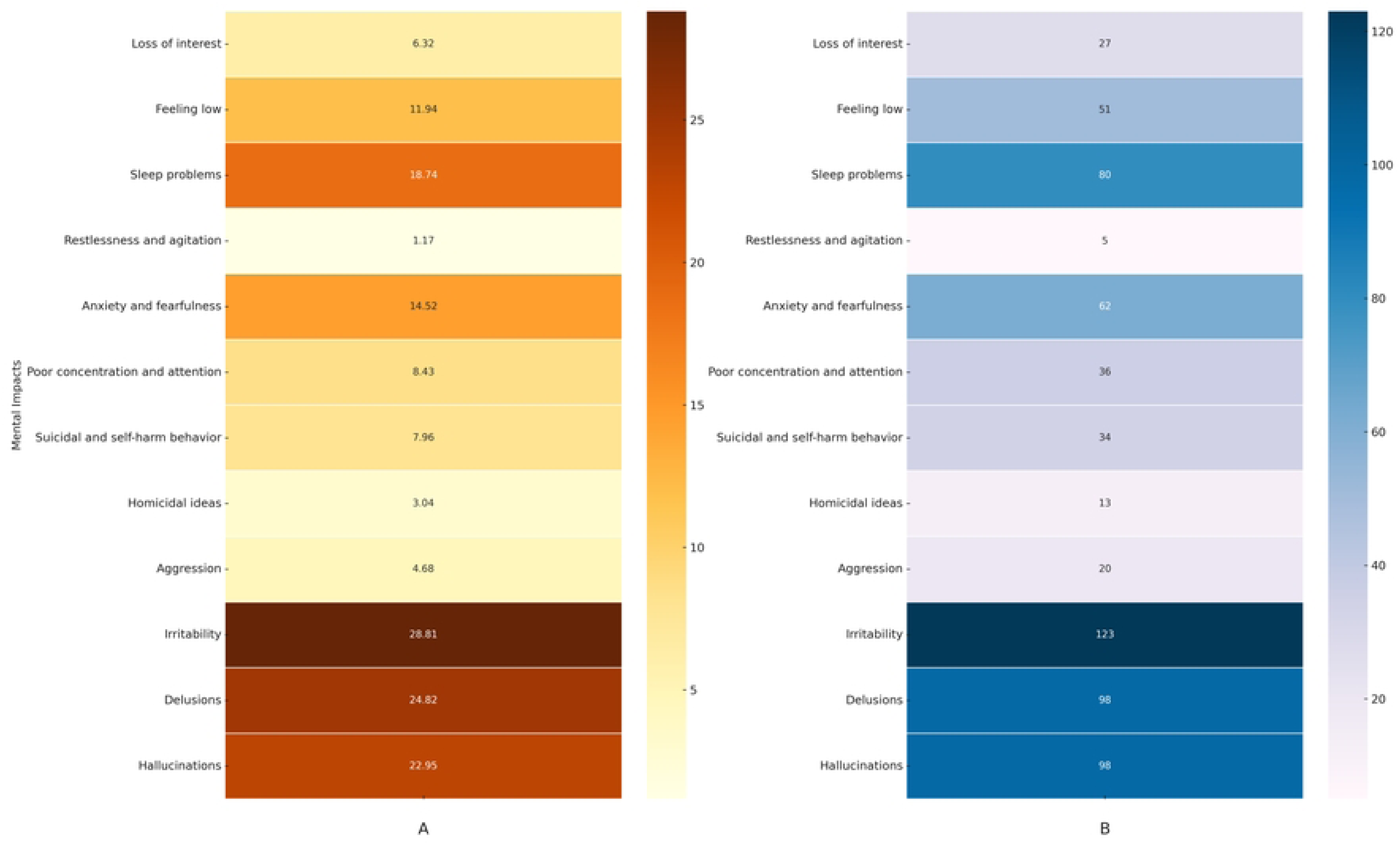
Heatmaps illustrating self-reported psychological/mental impacts due to Methamphetamine addiction. *(A) Percentage heatmap: Illustrates the relative percentages of self-reported psychological/mental impacts due to Methamphetamine addiction, with darker shades indicating a higher percentage of respondents experiencing each impact.* *(B) Frequency heatmap: Illustrates the absolute number of responders’ self-reported psychological/mental impacts due to Methamphetamine addiction, with darker shades indicating a higher number of respondents experiencing each impact.*

### Social Impact of Methamphetamine

Figure 3. illustrates the self-reported social impacts of Methamphetamine addiction. Interpersonal relationship problems and conflicts were the most frequently reported social impact, affecting 50.82% of participants. Financial problems were also common, reported by 32.08%, while stigmatization and social isolation were noted by 28.34%. Employment disruption impacted 10.07%, and legal problems were reported by 8.90%. Poor role performance affected 7.96%, and academic difficulties were the least reported, affecting only 2.58% of participants.

**Figure 3.**
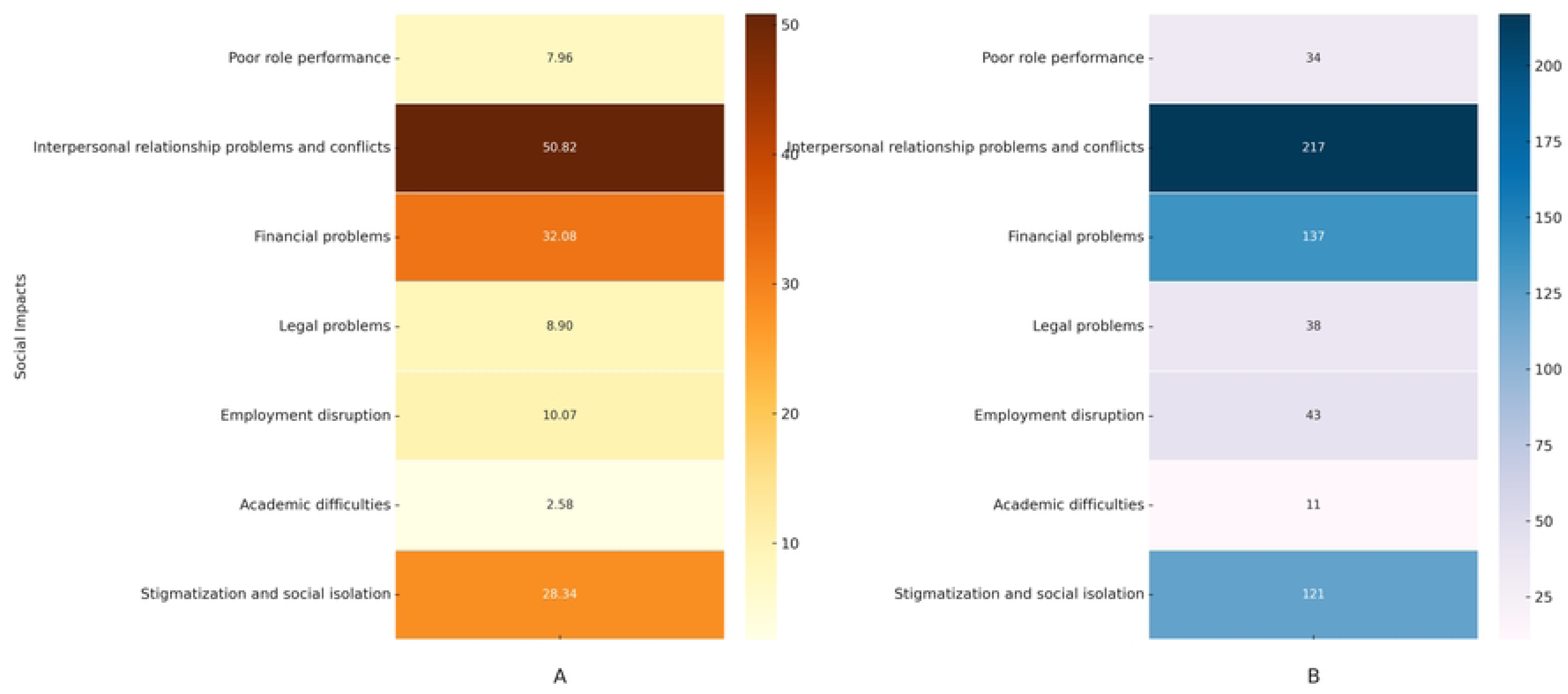
Heatmaps illustrating self-reported social impacts due to Methamphetamine addiction. *(A) Percentage heatmap: Illustrates the relative percentages of self-reported social impacts due to Methamphetamine addiction, with darker shades indicating a higher percentage of respondents experiencing each impact.* *(B) Frequency heatmap: Illustrates the absolute number of responders’ self-reported social impacts due to Methamphetamine addiction, with darker shades indicating a higher number of respondents experiencing each impact*.

## Discussion

The current study is, to the best of our knowledge investigating Methamphetamine addiction and its impact on adult individuals receiving treatment in a local context. By examining various physical, psychological, and social impacts of Methamphetamine addiction, the study contributes new insights into the complexities of addiction within this local context.

As revealed in the present study, the distribution of participants indicates a marked concentration in the Western Province, followed by North Western Province may reflect a regional pattern of Methamphetamine use and availability. The majority residing in urban and suburban areas suggest that MA addiction may be more commonly identified or reported in these settings, possibly due to higher drug availability. According to the Drug Related Statistics -2019 in Sri Lanka [27]The majority of drug-related arrests were reported from the Western Province, North Western Province, and Southern Province of Sri Lanka. These findings highlight the need for broader surveillance and the necessity of prompt preventive strategies.

The male predominance found in the present study is aligned with existing literature, denoting that men are more likely to engage in high-risk substance use behaviors, including Methamphetamine use [5,28]. Various cultural norms, social influences, and biological susceptibility perhaps contributed to this gender imbalance. In Sri Lankan culture, children and women are generally well protected within the family context, and substance use among women is strongly stigmatized by society’s behaviors [5]. This societal disapproval may contribute to the notably low prevalence of substance use among females. Consistent with previous studies [5], most of the study participants were aged between 18-30 years, and this highlights the vulnerability of young adults to use Methamphetamine. Adulthood often involves significant transitions, including entering the workforce and pursuing higher education generates stress and increases susceptibility to substance use [29]. These findings emphasize the importance of targeted preventive measures for young populations. The diversity in ethnic groups in the study sample implies that Methamphetamine addiction transcends demographic boundaries. While this distribution partly reflects the local population, it also highlights the necessity of implementing culturally competent preventive measures to protect young adults from Methamphetamine addiction. Nearly two-fifths of the participants reported being single, while a considerable number of participants were separated from their spouse. It also implies the social impact of Methamphetamine addiction due to its influence on splitting family relationships [30]. Compared with the findings of the National Dangerous Drug Control Board report in 2022 [31], more Methamphetamine-addicted individuals in the present study were separated (66 vs 12), and this highlights that Methamphetamine detrimentally influences breaking family relationships.

## Addiction to Methamphetamine

The findings on Methamphetamine addiction severity and patterns of use among participants reveal critical insights into the prevalence, onset, frequency, and influencing factors of Methamphetamine use. As revealed in the present study, most of the participants had moderate levels of Methamphetamine addiction, followed by 32.3% classified as severe and 29% as mild. This distribution suggests a significant proportion of users have progressed beyond mild addiction, highlighting the need for intensive intervention and rehabilitation programs tailored to different addiction severities. The present study findings demonstrated a high frequency of Methamphetamine consumption. Nearly half reported use on several days per week, while 28.1% consumed it daily. This pattern suggests a high dependency risk, particularly among daily users. Additionally, more than half reported using Methamphetamine within the past week. Prolonged Methamphetamine use causes disorders [32]. According to the Morbidity and Mortality Weekly Report published in the USA, 50% of persons using Methamphetamine in the past year met diagnostic criteria for past-year MA use disorder [28]. As is linear with previous findings, smoking and snorting are the most common methods of Methamphetamine consumption in the present study. Smoking and snorting allow for rapid drug absorption, leading to immediate and intense effects, which may contribute to higher addiction potential [33].

As revealed in the present study, the majority of participants were introduced to Methamphetamine by friends, followed by relatives. This finding highlights the strong role of peer influence in substance addiction, emphasizing the need for peer-focused prevention programs that address risky behaviors in social circles [34]. The accessibility of Methamphetamine is a considerable concern in the present study. As found a significant proportion of participants reported that Methamphetamine was fairly easy to access, while 36.5% found it easily accessible. In contrast, only a small percentage (5.6%) considered it difficult to obtain. The high availability of Methamphetamine in the community is a major concern, as easier access increases the likelihood of continued use and relapse [35]. These findings suggest the need for stronger regulatory measures, law enforcement efforts, and community interventions to restrict the availability and distribution of illicit drugs.

## Self-reported reasons for MA Addiction

The findings on self-reported reasons for Methamphetamine addiction indicate that peer influence is the most significant factor, with nearly half of the participants citing peer pressure as the reason for their drug use. This highlights the strong role of social circles in substance initiation and suggests that prevention strategies should focus on peer-led education and awareness programs. Additionally, 23.4% of participants reported that their peers were involved in ICE-related business, which could indicate exposure to drug distribution networks that encourage or normalize Methamphetamine use. Family-related factors such as isolation from family, lack of family closeness, and lack of parental support were reported by a smaller proportion of participants. Similarly, only 1.9% reported having parents who were drug users, and 1.2% mentioned experiencing excessive punishment as a reason for their addiction. These findings suggest that while family dynamics may influence drug use, they are not the primary drivers of Methamphetamine addiction in this study population [36]. Work-related reasons for Methamphetamine use were reported by a small but notable percentage of participants. Approximately 13% used Methamphetamine to maintain attention and concentration at work, while 10.5% reported using it to enhance job productivity. The association between substance use and work performance indicates the need for workplace mental health programs and stress management interventions to reduce reliance on drugs for job-related demands.

## Polydrug use

Present findings indicate that while a small portion of participants exclusively use Methamphetamine, the vast majority engage in polydrug use. This high rate of concurrent substance use underscores the complex nature of addiction, suggesting that individuals may use multiple drugs to achieve specific effects or to self-medicate varying psychological or physical needs. Within this polydrug subgroup, notable substances commonly used alongside methamphetamine include alcohol, cannabis, heroin, and tobacco, with a considerable percentage also reporting the use of other unspecified drugs. As revealed in a case series study in the local context, young adults consumed a mixture of substances including alcohol, heroin, cannabis, and amphetamines [4]. As found in previous studies, poly drug use was associated with rule-breaking behavior [29], violent and traumatic behavior, physical and sexual abuse [4], and various other psychiatric disorders such as major depression, post-traumatic stress disorder, panic attack, obsessive-compulsive disorder, and antisocial personality [37]. As cited by [4], compared with mature adult brains, young adults are more vulnerable to drug-seeking behaviors due to the “pleasure-seeking behavior” of the young, immature brains [38]. As revealed by Zolfaghari [39], prolonged consumption of Methamphetamine has a significant influence on the brain areas responsible for working memory, motor function, attention, visual interpretation, and speech processing.

## Physiological impact of MA addiction

The findings highlight a range of self-reported physical impacts of Methamphetamine use, demonstrating its potential to affect multiple bodily systems. The most frequently reported effects, including weight loss and loss of appetite, reflect profound effects of Methamphetamine which can lead to significant nutritional deficiencies over time. Also, dental problems were reported among participants. Other symptoms, such as malaise, chest pain, cough, and excessive sweating, further demonstrate the extent of the influence of MA addiction on the human body. Less common but noteworthy impacts including hair loss. As reported by Darke [22] addiction causes physical harm, including toxicity and mortality, cardiovascular/cerebrovascular pathology, dependence, and blood-borne virus transmission.

## Psychological impacts

Previous studies have documented the psychological harm of Methamphetamine addiction, including psychosis, depression, suicide, anxiety, and violent behaviors [22]. The present study findings underscore the breadth and severity of psychological impacts of Methamphetamine use, offering insights into both short-term disturbances and long-term mental health challenges. Irritability emerged as the most prevalent symptom, reflecting a heightened tendency toward emotional instability and which can lead to the disruption of interpersonal relationships and potentially escalate to aggression or conflict.[40–42]. High rates of delusions and hallucinations show the drug’s strong association with psychotic symptoms, which can emerge or intensify with frequent or prolonged use. Similarly, many studies have reported hallucinations as a frequently found psychotic symptom of chronic Methamphetamine use [42,43]. Comparatively, to previous studies, sleep problems are found to a considerable extent in the present study found in this study which indicates that Methamphetamine use can significantly disrupt normal sleep-wake cycles [38,44], contributing to fatigue, impaired judgment, and exacerbated mood swings [38]. Anxiety is more common among Methamphetamine chronic users [38,42,45,46]. High levels of anxiety/ fearfulness and feeling low found in the present study highlight the drug’s ability to influence the emotional and psychological well-being of users. These issues, tied with poor concentration and attention [46] can have profound implications for daily functioning and quality of life, often interfering with work [47]and education [48]. Of particular concern, suicidal thoughts or self-harm behaviors Mckertin [43] were reported by 7.96% of participants. Although homicidal ideas, aggression, and loss of interest were comparatively lower, these rates still signify a substantial risk that emphasizes the need for comprehensive mental health screenings and providing necessary treatments.

## Social impact of Methamphetamine addiction

The most prominent social impact reported was interpersonal relationship problems and conflict affecting over half of the participants. This finding underscores that Methamphetamine use can exert on personal connections, contributing to family disputes, breakdowns in friendships, and overall social dysfunction. Financial problems emerged as another key challenge, highlighting the economic toll associated with drug expenses and potential job loss or reduced productivity. Additionally, stigmatization and social isolation reflect the broader social barriers and negative attitudes that individuals with substance use disorders often face, which can hinder access to support and worsen mental health outcomes. Although employment disruption (10.07%, legal problems (8.9%), poor performance, and academic difficulties were less frequently reported, they still indicate that Methamphetamine addiction can detrimentally affect nearly every aspect of daily life, from maintaining steady employment to fulfilling societal and personal responsibilities. As reported previously, chronic use of Methamphetamine causes social isolation due to social withdrawal [20].

## Limitations

The present study has several limitations. The descriptive cross-sectional design employed in this study only captures data at one point in time, thus it prevents determining cause-and-effect relationships between variables. Data were collected through an interviewer-administered questionnaire and it may influence for responses of the participants. Further, true and honest responses of the participants are difficult except to a greater extent, due to social desirability bias inherent in questionnaire-based studies.

## Conclusions

This study sheds light on the growing public health concern of Methamphetamine addiction is Sri Lanka, particularly among young adult males. The findings reveal the high prevalence of moderate to severe addiction, influenced predominantly by peer pressure and facilitated by easy access to Methamphetamine. The widespread occurrence of polydrug use and the physical, psychological, and social repercussions, including disrupted family relationships and workplace challenges, highlight the complexity of addiction. The study bridges a significant research gap by providing localized insights into patterns and consequences of MA addiction. Thus, targeted peer-led and culturally sensitive prevention strategies, especially for young adults, are crucial. Strengthening community-based mental health and rehabilitation services, together with strict control over drug accessibility, will be vital to address this rising issue effectively.

## Data Availability

All relevant data are within the manuscript and its Supporting Information files.

## Acknowledgments

The authors would like to thank all the participants and the NIMH clinical staff and administration officers.

## Author contributions

Conceptualization: K.A. Sriyani, N.A.A.I. Nishshanka, T.N.L. Samarathunga, S.W. Inoka, R. Suharna

Data curation: , D.K.M. De Silva, N.A.A.I. Nishshanka Formal analysis: D.K.M. De Silva, N.A.A.I. Nishshanka

Investigation: N.A.A.I. Nishshanka, T.N.L. Samarathunga, S.W. Inoka, R. Suharna

Methodology: K.A.Sriyani, D.K.M. De Silva, N.A.A.I. Nishshanka, T.N.L. Samarathunga, S.W. Inoka, R. Suharna

Project administration: K.A.Sriyani

Writing – original draft: D.K.M. De Silva, K.A.Sriyani Writing – review & editing: D.K.M. De Silva, K.A.Sriyani

